# Mental health problems in the general population during and after the first lockdown phase due to the SARS-Cov-2 pandemic: Rapid review of multi-wave studies

**DOI:** 10.1101/2020.12.03.20243196

**Authors:** Dirk Richter, Steffi Riedel-Heller, Simeon Zuercher

**Affiliations:** Bern University Hospital for Mental Health, Center for Psychiatric Rehabilitation, Bern, Switzerland; University of Bern, Department of Psychiatry and Psychotherapy, Bern, Switzerland; Bern University of Applied Sciences, Department of Health Professions, Bern, Switzerland; University of Leipzig, Institute of Social Medicine, Occupational Health and Public Health (ISAP), Leipzig, Germany; Bern University Hospital for Mental Health, Center for Psychiatric Rehabilitation, Bern, Switzerland University of Bern, Department of Psychiatry and Psychotherapy, Bern, Switzerland

## Abstract

**Aims:** The SARS-Cov-2 pandemic and the lockdown response are assumed to have increased mental health problems in general populations compared to pre-pandemic times. The aim of this paper is to review studies on the course of mental health problems during and after the first lockdown phase.

**Methods:** We conducted a rapid review of multi-wave studies in general populations with time points during and after the first lockdown phase. Repeated cross-sectional and longitudinal studies that utilized validated instruments were included. The main outcome was whether indicators of mental health problems have changed during and after the first lockdown phase. The study was registered with PROSPERO No. CRD42020218640.

**Results:** 23 studies with 56 indicators were included in the qualitative review. Studies that reported data from pre-pandemic assessments through lockdown indicated an increase in mental health problems. During lockdown no uniform trend could be identified. After lockdown mental health problems decreased slightly.

**Conclusions:** As mental health care utilization indicators and data on suicides do not suggest an increase in demand during the first lockdown phase, we regard the increase in mental health problems as general distress that is to be expected during a global health crisis. Several methodological, pandemic-related, response-related and health policy-related factors need to be considered when trying to gain a broader perspective on the impact of the first wave of the pandemic and the first phase of lockdown on general populations’ mental health.

---

The SARS-Cov-2 pandemic has affected nearly the entire world and has led to considerable loss of life and high rates of physical morbidity. Moreover, the pandemic has seriously affected economies and individual livelihoods. Previous epidemics had tremendous negative consequences on the mental health of various population groups such as health care workers and survivors of the infectious disease (Zürcher et al., 2020). However, past epidemics have also negatively impacted the mental health of general populations at large (Zürcher et al., 2020). Therefore, during the first wave of the Coronavirus pandemic and the first phase of lockdowns, there were widespread fears concerning mental health problems beyond population groups that were directly affected by the illness (The Lancet Infectious Diseases, 2020). Some professional societies even feared a ‘tsunami of mental illness’ (Royal College of Psychiatrists, 2020).

Several systematic reviews and meta-analyses have analysed the extent of mental health problems in the general population during the lockdown phase in spring/summer 2020. Pooled prevalence rates for depression, anxiety, and distress reached 30 to 40 percent (Krishnamoorthy et al., 2020, Luo et al., 2020, Salari et al., 2020). Although these reviews did not directly compare pre-pandemic and pandemic time points, the prevalence rates suggest an increase in mental health problems during the first months in spring and early summer 2020 compared to pre-pandemic assessments. These prevalence rates, however, should to be interpreted with some caution due to methodological and psychopathological issues (Riedel-Heller and Richter, 2020). To start with methodological caveats, many studies suffer from problems with sampling and sample size – which is understandable in the circumstances of an immediate outbreak. From a psychopathological perspective, it is unclear to what extent the prevalence rates that are measured with self-report instruments reflect common distress that is to be expected in such public health crises and to what extent this distress will result in increasing rates of mental disorders and health care utilization demand. Therefore, we have decided to remain cautious and to stick to the terminology of ‘mental health problems’ rather than ‘mental illness’.

Another point that needs to be considered is the research design of most studies in this field. Most studies have utilized either a one-time cross-sectional design or have compared pre-lockdown data to cross-sectional results from data collection during lockdown. As nearly all countries eased their restrictions during summer 2020 and many have already entered the second phase of strict non-pharmacological interventions, we seek to explore the course of mental health problems during and after the first phase of lockdowns that is closely related to the first pandemic wave. A longitudinal perspective can help to solve some of the methodological and psychopathological problems and may inform about possible future developments concerning mental health during the pandemic.

## Methods

We have conducted a rapid review of multi-wave studies that gathered data from general populations during and/or after the first lockdown phase in 2020. Rapid reviews are recommended in cases where swift information is needed in order to inform policies and administrative responses to health-related challenges (Tricco et al., 2017). In doing so, rapid reviews waive some characteristics of systematic reviews to facilitate a publication that will be utilized more quickly than is common with systematic review. Our rapid review was registered with PROSPERO CRD42020218640.

We searched Pubmed (including preprint servers medRxiv, bioRxiv, arXiv, Research Square, and SSRN), PsychInfo and the preprint server PsyArXiv with search terms that have been adapted to the requirements of each database (see details in appendix). We also searched the search machine Google Scholar with very broad search terms. Inclusion criteria were as follows: studies that covered at least two time points during the first lockdown phase or at least one time point during lockdown and one time point after easing of public health restrictions. Studies that, for whatever reason, reported additional pre-pandemic data were not excluded. Both, repeated cross-sectional surveys and longitudinal panel studies were deemed to be analysed. Included languages were English, French, Dutch, Spanish and German. Publications in other languages were excluded. Further, we included only studies that utilized psychometrically validated instruments for assessing mental health problems. Any reporting modus (means, prevalence rates, regression coefficients) was included as long as data on at least two time points were reported. Exclusion criteria were a) health care workers, b) survivors or patients with SARS-CoV-2, c) vulnerable populations with a risk of being marginalized, that live in precarious situations, with poor access to health care services, with chronic physical conditions (e.g. homeless people, people with pre-existing mental illness, indigenous populations, cancer patients), d) specific subpopulations (e.g. students, young adults, seniors). The search was conducted by DR and randomly checked by SZ.

We extracted the following data: authors, country of data collection, publication status (preprint vs. peer-reviewed publication), sampling procedure, sample size, utilized assessment instrument, scale means or prevalence rates at the following time points: pre-lockdown, first and last timepoints during lockdown, post-lockdown. Results were briefly summarized in a separate column. In the case of non-reporting of means or prevalence data (e.g. when coefficients were reported), we extracted the result as provided by the authors. When only figures were presented, we estimated the exact numbers by measuring the bars. In cases where prevalence rates and scores were reported from the same study, we extracted both indicators. In order to provide a simplified overview on the changes during and after the first lockdown phase, we will display the results in three figures in the appendix. Data extraction was conducted by DR and checked by SZ. Quality appraisal was conducted with an adapted instrument (Munn et al., 2015). Items were rated on the options: yes, no, unclear. Following criteria were covered: sampling frame, sampling method, sample size, subjects and setting description, coverage, standardized procedures, and response rate. Quality appraisal was conducted by SZ and randomly checked by DR. The application of meta-analytical methods was impossible due to the heterogeneity of instruments, timepoints and measures. There was no funding source for this study.

## Results

After study selection (Figure 1), we included 23 publications into the qualitative and narrative synthesis. Four of these studies covered the USA, another four were from Germany (one jointly with Austria) and three were from the United Kingdom. Apart from two publications from China, the remainder of the studies were single publications from various countries that predominantly covered European populations. 16 studies were peer-reviewed, 7 were published as preprints.

**Figure 1:**
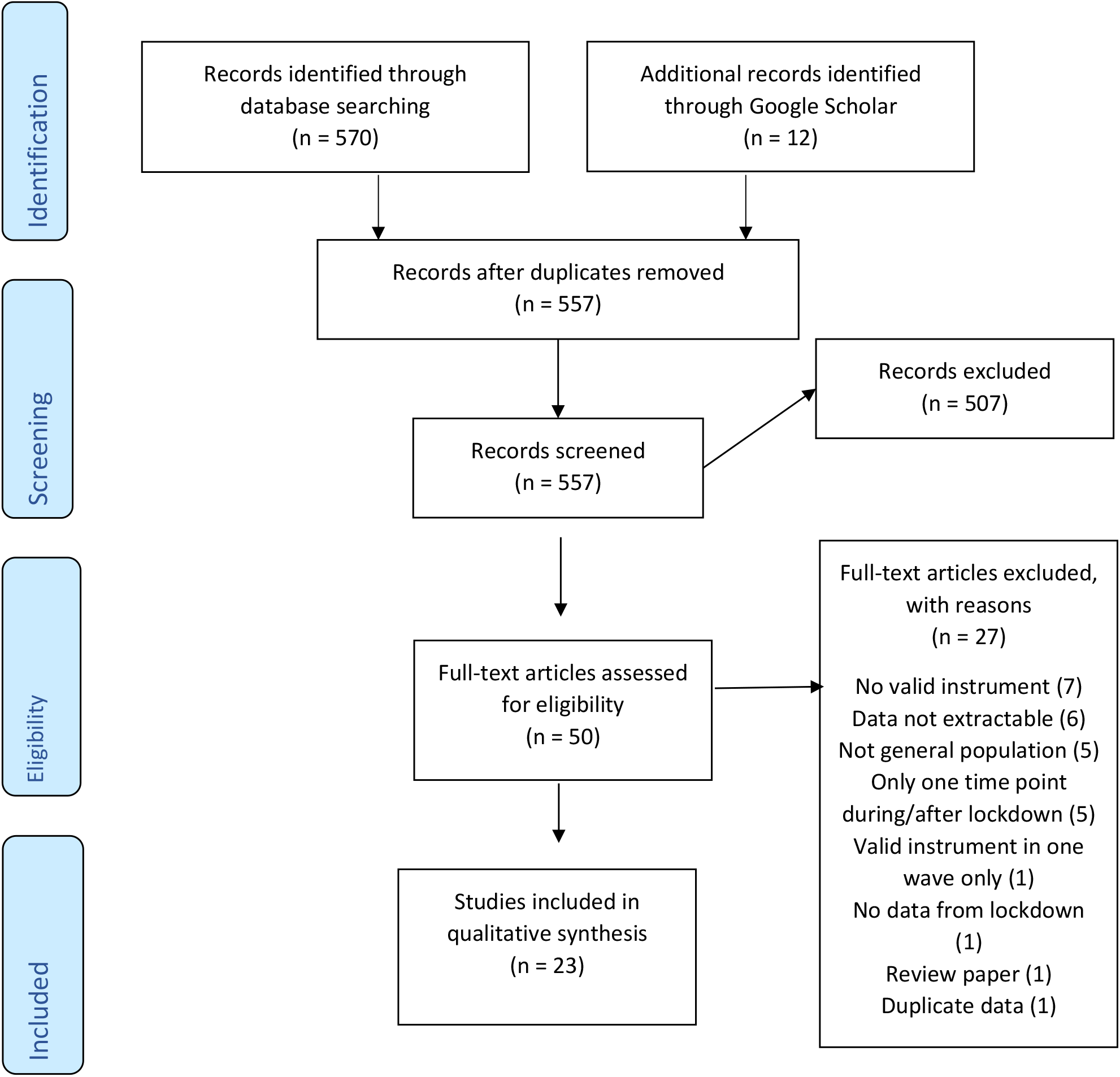
Flow-Chart according to PRISMA

We identified the following study characteristics (Table 1). The sample sizes ranged from less than 200 to 90,000 per time point. 12 studies managed to collect data from representative samples while 11 utilized convenience or snowball sampling. The included studies reported results from 56 indicators. This means that many studies utilized two or three instruments simultaneously (e.g. anxiety plus depression). 7 studies reported data from pre-pandemic time points, 6 from post-lockdown phase. The instrument most utilized was the ‘Depression, Anxiety, and Stress Scales’, followed by the ‘Patient Health Questionnaire’ and the ‘Generalized Anxiety Disorder Assessment’. Different versions of these and other scales were used. Our quality appraisal (Appendix) has shown that many studies have issues with sampling, recruitment and coverage, while the research procedures and the description of subjects and settings were well reported.

**Table 1:** Characteristics of multi-wave studies reporting on mental health problems for at least two time points (during the first lockdown/pandemic wave or during the first lockdown/pandemic wave and a time point after easing of public health restrictions)

| Author | Country | Pre-print (PP) / Peer-reviewed (PR) | Sampling | Sample size | Instrument | Pre-Lockdown | First Lockdown | Latest Lockdown | Post-Lockdown | Results (effect size if provided by original studies) |
| --- | --- | --- | --- | --- | --- | --- | --- | --- | --- | --- |
| Canet-Juric et al.(Canet-Juric et al., 2020) | Argentina | PR | Non-representative Online Survey, Social networks | 6,057 | BDI-II (score) | .. | 8.74 | 9.41 | .. | Significant increase; small effect size |
|  |  |  |  |  | STAI (score) | .. | 1.16 | 1.11 | .. | Significant decrease; small effect size |
|  |  |  |  |  | PANAS (neg.) (score) | .. | 17.6 | 17.14 | .. | Significant decrease; small effect size |
|  |  |  |  |  | PANAS (pos.) (score) | .. | 24.06 | 23.79 | .. | Significant decrease; small effect size |
| Chandola et al.(Chandola et al., 2020) | United Kingdom | PP | Representative Household Panel Survey | 10,266 to 11,582 | GHQ-12 (prevalence) | 24 | 37.2 | 33.8 | .. | Significant increase from pre-lockdown to first Lockdown; decrease first lockdown to latest lockdown (no formal significance test) |
| Daly et al.(Daly et al., 2021) | United States | PR | Representative Online Survey | 5,075 to 6,819 | PHQ-2 (prevalence) | 8.9 | 10.5 | 14.2 | .. | Significant increase across all timepoints |
|  |  |  |  |  | PHQ-2 (score) | 0.69 | 0.82 | 1.08 | .. | Significant increase across all timepoints |
| De Quervain et al.(de Quervain et al., 2020) | Switzerland | PP | Non-representative online Survey | 5,120 to 10,438 | PHQ-9 (prevalence) | 3.4 | 9.1 | 12.5 | .. | Significant increase across all timepoints |
| Duan et al.(Duan et al., 2020) | China | PR | Random online survey | 1,390 | PSS-10 (score) | .. | 2.56 | 2.36 | .. | Significant decrease; medium effect size |
| Du Roscoät(du Roscoät, 2020) | France | PP | Representative online survey | 2,000 | HADS (Anx.) (prevalence) | .. | 26.7 | 18.4 | 16.9 | Significant decrease from first lockdown to post-lockdown |
|  |  |  |  |  | HADS (Depr.)<br>(prevalence) | .. | 19.9 | 18.6 | 12.1 | Significant decrease from first lockdown to post-lockdown |
| Fancourt et al. (Fancourt et al., 2020) | United Kingdom | PP | Representative online survey | 36,520 | PHQ-9 (score) | .. | ≈7.36 | ≈5.96 | ≈5.56 | Significant decrease from first lockdown to post-lockdown |
|  |  |  |  |  | GAD-7 (score) | .. | ≈5.92 | ≈4.44 | ≈4.04 | Significant decrease from first lockdown to post-lockdown |
| Filgueiras & Stults-Kolehmainen (Filgueiras and Stults-Kolehmainen, 2020) | Brazil | PP | Non-representative online survey | 360 | PSS-10 (score) | .. | 20.54 | 22.03 | .. | Significant increase |
|  |  |  |  |  | PSS-10 (prevalence) | .. | 65.8 | 74.7 | .. |  |
|  |  |  |  |  | FDI (score) | .. | 65.32 | 70.31 | .. | Significant increase |
|  |  |  |  |  | FDI (prevalence) | .. | 62.2 | 72.2 | .. |  |
|  |  |  |  |  | STAI (score) | .. | 43.61 | 49.88 | .. | Significant increase |
|  |  |  |  |  | STAI (prevalence) | .. | 48.2 | 64.3 | .. |  |
| Fiorillo et al. (Fiorillo et al., 2020) | Italy | PR | Non-representative online survey | 20,720 | DASS-21 (Depr.) (score) | .. | 12.1 | 13.1 | .. | Significant increase |
|  |  |  |  |  | DASS-21 (Anx.) (score) | .. | 7.5 | 8.5 | .. | Significant increase |
|  |  |  |  |  | DASS-21 (Stre.) (score) | .. | 16 | 17.2 | .. | Significant increase |
| Gopal et al. (Gopal et al., 2020) | India | PR | Non-representative online survey | 159 | GAD-7 (prevalence) | .. | 10.4 | 12.7 | .. | Significant increase |
|  |  |  |  |  | PHQ-2 (prevalence) | .. | 14.8 | 26.1 | .. | Significant increase |
| Hetkamp et al.(Hetkamp et al., 2020) | Germany | PR | Non-representative online survey | 16,245 | GAD-7 (prevalence) | 7.2 | 37 | 22.1 | .. | Increase pre-lockdown to first lockdown; decrease from first lockdown to latest lockdown (no test statistic provided) |
| Holman et al.(Holman et al., 2020) | United States | PR | Representative online survey | 2,122 to 2,234 | ASDS-5 (score) | .. | ≈1.75 | ≈1.95 | .. | Significant increase |
|  |  |  |  |  | BSI (score) | .. | ≈0.55 | ≈0.76 | .. | Significant increase |
| Kikuchi et al.(Kikuchi et al., 2020) | Japan | PR | Non-representative online survey | 2,078 | K6 (prevalence) | .. | 9.34 | 11.31 | .. | Significant increase |
|  |  |  |  |  | K6 (score) | .. | 4.79 | 5.6 | .. | Significant increase |
| Kimhi et al.(Kimhi et al., 2020) | Israel | PR | Representative online survey | 300 | BSI (score) | .. | 2.35 | .. | 2.19 | Significant decrease |
| Mata et al.(Mata et al., 2020) | Germany | PP | Representative online survey | 3,500 | STAI (score) | .. | 1.8 | 1.7 | 1.66 | Significant decrease between first lockdown and latest lockdown/post-lockdown |
|  |  |  |  |  | PHQ-2 (score) | .. | .. | 1.5 | 1.43 | Significant decrease between first lockdown and latest lockdown/post-lockdown |
| O'Connor et al.(O'Connor et al., 2020) | United Kingdom | PR | Representative online survey | 2,604 to 3,077 | PHQ-9 (prevalence) | .. | 26.1 | 23.7 | .. | Non-significant decrease |
|  |  |  |  |  | GAD-7 (prevalence) | .. | 21 | 16.8 | .. | Significant decrease |
| Planchuelo-Gómez et al.(Planchuelo-Gómez et al., 2020) | Spain | PP | Non-representative online survey | 1,174 to 3,550 | DASS-21 (Anx.) (prevalence) | .. | 32.45 | 37.22 | .. | Significant increase |
|  |  |  |  |  | DASS-21 (Anx.) (score) | .. | 3.15 | 3.6 | .. | Significant increase |
|  |  |  |  |  | DASS-21 (Depr.) (prevalence) | .. | 44.11 | 46.42 | .. | Significant increase |
|  |  |  |  |  | DASS-21 (Depr.) (score) | .. | 5.06 | 5.55 | .. | Significant increase |
|  |  |  |  |  | DASS-21 (Stre.) (prevalence) | .. | 37 | 49.66 | .. | Significant increase |
|  |  |  |  |  | DASS-21 (Stre.) (score) | .. | 6.5 | 7.94 | .. | Significant increase |
| Schnell & Krampe(Schnell and Krampe, 2020) | Germany and Austria | PR | Non-representative online survey | 1,527 | PHQ-4 (score) | .. | 3.21 | .. | 3.87 | Significant increase |
|  |  |  |  |  | PHQ-4 (prevalence) | .. | 42 | .. | 40 | No significant decrease |
| Staples et al.(Staples et al., 2020) | Australia | PR | Non-representative online survey | 5,454 | K-10 (score) | 31.2 | 31.4 | 30.7 | .. | Non-significant changes |
|  |  |  |  |  | PHQ-9 (score) | 14.3 | 14.4 | 14.1 | .. | Non-significant changes |
|  |  |  |  |  | GAD-7 (score) | 12.1 | 12.5 | 11.9 | .. | Significant increase between pre-lockdown and first lockdown; significant decrease between first lockdown and latest lockdown (small effect) |
| Twenge & Joiner(Twenge and Joiner, 2020) | United States | PR | Representative online survey | 17,067 to 90,798 | PHQ-2 (prevalence) | 6.6 | 23.5 | 24.9 | .. | Significant increase between pre-lockdown and first lockdown; significant increase between first lockdown and latest lockdown |
|  |  |  |  |  | GAD-2 (prevalence) | 8.2 | 30.8 | 29.4 | .. | Significant increase between pre-lockdown and first lockdown; significant decrease between first lockdown and latest lockdown |
| Wang et al.(Wang et al., 2020) | China | PR | Non-representative online survey | 861 to 1,304 | IES-R (score) | .. | 32.98 | 30.76 | .. | Significant decrease |
|  |  |  |  |  | DASS-21 (Stre.) (score) | .. | 7.76 | 7.86 | .. | Non-significant increase |
|  |  |  |  |  | DASS-21 (Anx.) (score) | .. | 6.16 | 6.15 | .. | Non-significant decrease |
|  |  |  |  |  | DASS-21 (Depr.) (score) | .. | 6.25 | 6.38 | .. | Non-significant increase |
| Zacher & Rudolf(Zacher and Rudolph, 2020) | Germany | PR | Representative online survey | 979 | PANAS (pos.) (score) | 4.49 | 4.46 | 4.37 | 4.28 | Non-significant decrease between pre-lockdown to first lockdown; Significant decrease after first lockdown |
|  |  |  |  |  | PANAS (neg.) (score) | 2.64 | 2.62 | 2.56 | 2.49 | Non-significant decrease between pre-lockdown to first lockdown; Significant decrease after first lockdown |
| Zhou et al.(Zhou et al., 2020) | United States | PR | Representative online survey | 442 to 1,021 | DASS-21 (Stre.) (score) | .. | 7.39 | 6.13 | .. | Significant decrease |
|  |  |  |  |  | DASS-21 (Anx.) (score) | .. | 5.77 | 4.36 | .. | Significant decrease |
|  |  |  |  |  | DASS-21 (Depr.) (score) | .. | 7.14 | 5.69 | .. | Significant decrease |

≈, results provided in graphical format only
ASDS-5, Acute Stress Disorder Scale-5; BDI-II, Beck Depression Inventory-II; BSI, Brief Symptom Inventory; DASS-21 (Anx.), Depression, Anxiety, Stress Scale - Anxiety; DASS-21 (Depr.), Depression, Anxiety, Stress Scale - Depression; DASS-21 (Stre.), Depression, Anxiety, Stress Scale - Stress; FDI, Filgueiras Depression Inventory; GAD-2, Generalised Anxiety Disorder-2; GAD-7, Generalised Anxiety Disorder-7; GAD-7, Generalized Anxiety Disorder-7; GHQ-12, 12-item General Health Questionnaire; HADS (Anx.), Hospital Anxiety and Depression Scale; HADS (Depr.), Hospital Anxiety and Depression Scale; IES-R, Impact of Event Scale-Revised; K-10, Kessler Psychological Distress Scale; K6, Kessler Psychological Distress Scale; NA, text; PANAS (pos.), Positive and Negative Affect Schedule -Positive Affect; PHQ-2, Patient Health Questionnaire-2; PHQ-4, Patient Health Questionnaire-4; PHQ-9, Patient Health Questionnaire-9; PSS-10, Perceived Stress Scale-10; STAI, State-Trait Anxiety Inventory

The changes of means, prevalence rates and coefficients were not uniform across indicators, types of mental health problems and time points (see Figures 2-4, Appendix). The general impression was as follows: a huge variation during lockdown but no uniform trends and a slight decrease of indicator data after lockdown. We also noted an increase from pre-pandemic times to lockdown.

We also looked for trends in specific types of mental health problems (e.g. anxiety or depression) and for trends in countries with more than two studies. Again, we found no uniform trends across studies.

## Discussion

This rapid review has compiled data from multi-wave studies that analysed mental health problems during and after the first lockdown phase of the SARS-Cov-2-pandemic. We did not find a uniform trend of mental health problems and assume – without being able to quantify – a considerable heterogeneity. However, the most likely ‘big picture’ that emerged is as follows: as suggested by previous systematic reviews (Krishnamoorthy et al., 2020, Luo et al., 2020, Salari et al., 2020), an increase in mental health problems can be seen from pre-pandemic time points to the first phase of the lockdown. During the first phase, we see a diversity of trends, some increasing, some decreasing and some with no changes. After the easing of the restrictions, we mainly find a slight decrease in mental health problems. This decrease, however, does not reach pre-pandemic levels.

Several methodological, psychopathological, pandemic-related, lockdown-related and policy-related issues have to be considered while interpreting this somewhat unclear picture.

1. Methodologically, we found a diversity of instruments, of versions of the same instrument and sampling approaches that were utilized in the studies. Additionally, we found many studies that used more than one indicator answered by the same respondents. This methodological challenge is known from previous reviews on prevalence changes of mental illness and needs to be accounted for as the effect sizes are dependent (Richter et al., 2019, Fernandez-Castilla et al., 2020).
2. In terms of psychopathology, we found several mental health problems that were addressed: anxiety, depression and distress among them prominently. While these are negative emotional reactions, it is not clear what kind of problem is most relevant and whether these problems reflect general distress. And again, we did not find a clear trend that emerged in this regard. Additionally, we have seen statistically relevant differences in means and prevalences between time points during and after lockdown that *prima facie* do not indicate large clinical relevance and effect size.
3. In many countries, the mental health-related consequences of the pandemic cannot be clearly separated from lockdown effects as lockdowns are commonly implemented when infection rates are high. Nevertheless, we assume that emotional reactions to the infection are a major background of the mental health problems that have emerged. Although consequences from financial hardship will certainly impact emotional states during the pandemic, research has shown that people who received financial support during the pandemic reported less mental health needs than those who did not receive such support (Berkowitz and Basu, 2020). Many countries, even in the developing world (e.g. Brazil) have supported their citizens with financial aid or furlough programs during the pandemic (Richter, 2021). In this regard, it needs to be considered that the first wave of the pandemic in larger countries did not hit the entire country at the same time. In many countries, the pandemic spread from hotspots (e.g. Lombardy in Italy or the Northeast of the United States) to other areas. Data from the United States Centers for Disease Control suggest that mental health problems are to a certain extent correlated to infection rates (CDC, 2020). Therefore, the timing of data collection and actual infection rates in the region where respondents are crucial information that is commonly missing in the publications. These methodological problems raise some doubts on nationwide surveys that cover differently affected regions, particularly in larger countries.
4. The implementation of non-pharmacological interventions, summarized as lockdown, was not uniform across countries or sometimes even within countries (e.g. the USA). Some states implemented very strict measures that locked citizens up in their houses, other states ordered curfews at night-time only, and other jurisdictions again adopted a ‘lighter’ approach (Oxford University, 2020). As the rigidity and time length of restrictions is known to be of importance in terms of mental health problems (Brooks et al., 2020, Huremovic, 2019), we assume that those differences need to be accounted for when interpreting data on mental health during and after lockdown. Further, the success of measures in terms of the reduction or even suppression of infection rates needs to be considered.
5. Alongside differences in the implementation of non-pharmacological intervention, states have also shown a diversity of policy responses. While some political leaders have clearly and consistently communicated the risks of the pandemic and of non-adherence to restrictions, others have denied the public health crisis and have defied recommendations from experts. As empirical research has demonstrated, this clearly had an impact on the general public. In the USA, for example, the politicization of the pandemic has led to diverse views and behavioural responses according to political camps (Zhao et al., 2020). We assume that these differences will also impact the emotional response to the pandemic. When people see the virus as non-existent or to be of minor risk, there is no reason to be worried about it. In addition, states have differed to a certain extent in the welfare response that aimed at mitigating the economic and psychosocial consequences of the pandemic. The longer the pandemic is not sufficiently suppressed, the more important the welfare state response in terms of mental health becomes.

As data on other mental health indicators suggest, the first lockdown phase has – in general – not led to an increase in mental health care utilization. A UK study reported that the demand for mental health care decreased partly due to fears of becoming infected in health care settings (Chen et al., 2020). A large German statutory health insurer published a report that indicated a sharp increase in mental health-related sick leave during the first pandemic peak that had returned to ‘normal’ levels during spring and summer of 2020. This report concluded: “It would be inappropriate to derive increasing mental illness rates from these data.” (Techniker Krankenkasse, 2020: 44; our translation) Also, suicide rates and suicide attempt rates in various countries do not seem to have risen compared to pre-pandemic times (e.g. Hernandez-Calle et al., 2020, John et al., 2020, Leske et al., 2020). Whether these trends will hold in further infection or lockdown phases, remains to be seen. Our assumption that post-lockdown mental health problems have not decreased to pre-pandemic levels, is concerning in this regard. Newly imposed restrictions during the second phase of lockdown together with infection rates that are much higher than during the first pandemic wave and increasing economic worries may induce more mental health problems in future weeks and months. This may also result in a higher demand for mental health services.

## Conclusions

We conclude from this rapid review that mental health problems in the general population have not essentially changed during the first lockdown after they have risen compared to pre-pandemic times. After easing of lockdown restrictions, they have decreased to a level that is assumingly higher than before the pandemic. As many data sources do not indicate an increasing demand for mental health care utilization during the first wave of the pandemic, we interpret these mental health problems generally as distress that is to be expected during a global public health crisis. This conclusion, however, does not disregard that some individuals or some population groups have suffered psychologically over and above the commonly to be expected distress in the first half of 2020. Again, studies on people with pre-existing mental disorders, for example, do not generally suggest worse outcomes during the first lockdown phase. While some studies see more distress on this group (O’Connor et al., 2020, Liu et al., 2020), others reject this hypothesis (Pinkham et al., 2020, Schutzwohl and Mergel, 2020). And our conclusion does not disregard that there is a certain risk for increasing mental illness and demand for mental health care in the general population the longer the pandemic and its economic and psychosocial consequences will last.

In terms of methodology, we caution against the over-interpretation of results from single studies on mental health problems during the pandemic. Similar to meta-analyses of clinical trials which oftentimes provide conflicting results on health care interventions, only aggregate and synthesized data are able to inform on general trends. This is particularly the case during this pandemic as various confounding factors need to be considered when trying to get a clearer perspective on the complexity of mental health problems in such a crisis.

## Data Availability

Data are available in the manuscript.

## Appendix: Search strategy

### Pubmed

(Covid-19 OR lockdown OR SARS-Cov-2) AND (mental OR psychiatr* OR psycholog*) AND (repeated OR longitudinal OR wave* OR during [TI])

### PsychInfo (via Ovid)

((Covid-19 or lockdown or SARS-Cov-2) and (mental or psychiatr* or psycholog*)).mp. and ((repeated or longitudinal or wave*).mp. or during.ti.) [mp=title, abstract, heading word, table of contents, key concepts, original title, tests & measures, mesh]

### PsyArxiv

(Covid-19 OR lockdown OR SARS-Cov-2) AND (mental OR psychiatr* OR psycholog*) AND (repeated OR longitudinal OR wave*) Restricted to 2020

**Appendix Figure 1:**
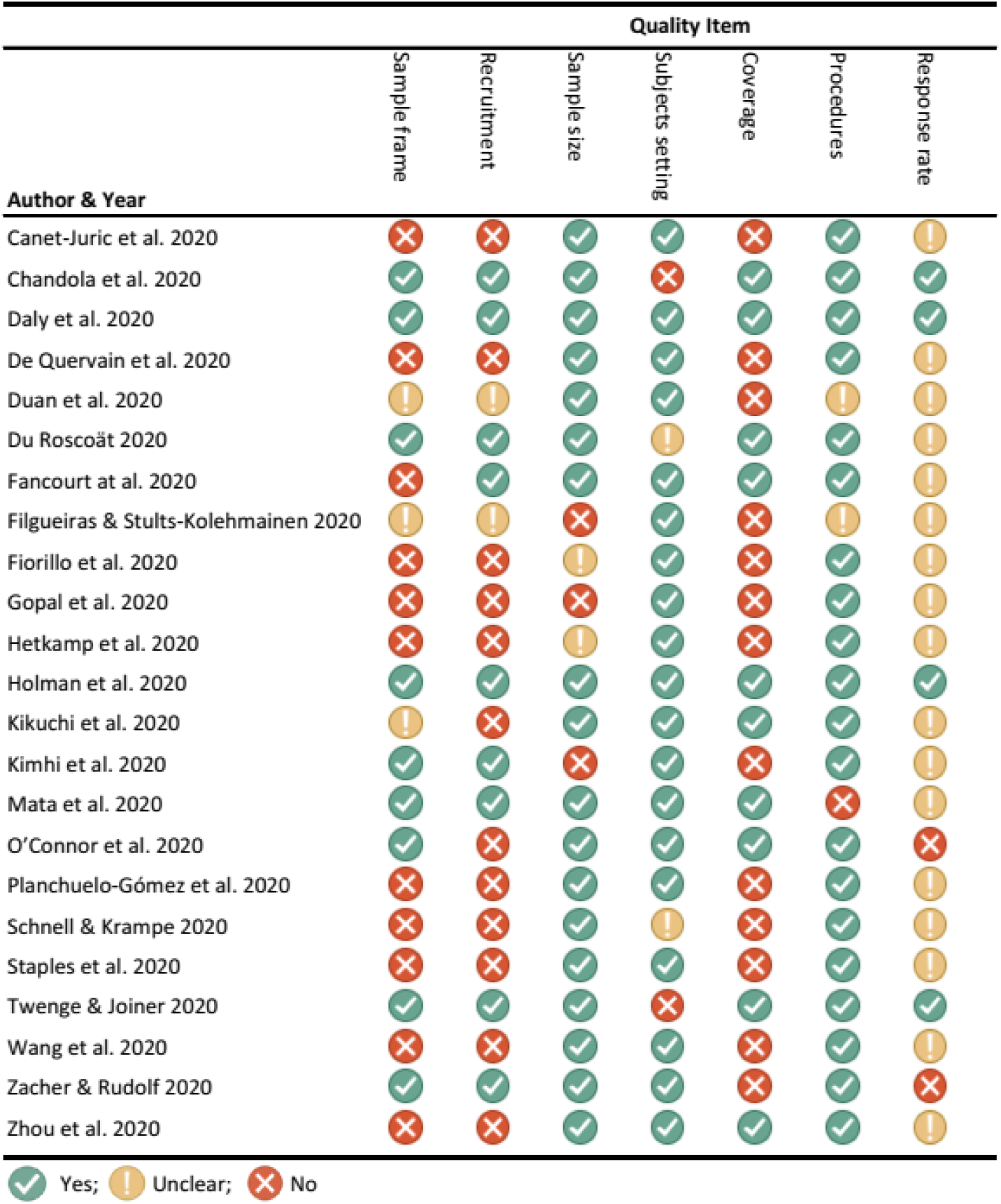
Quality Appraisal - Judgements about each methodological quality item for each included study assessed by an adapted version of the Joanna Briggs Institute Critical Appraisal (JBI) tools for Prevalence Studies

**Appendix Figure 2:**
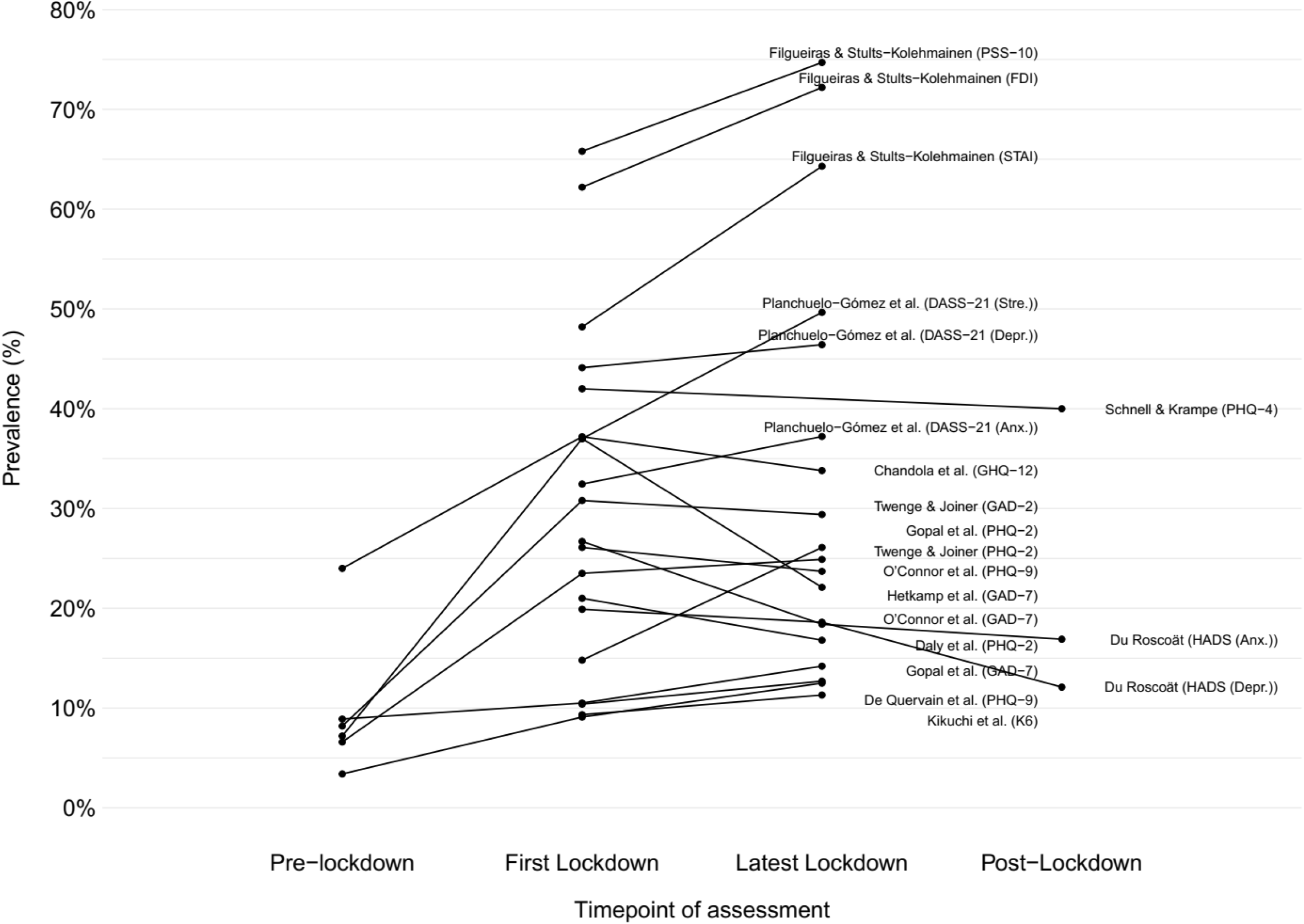
Prevalence rates of mental health problems during and after lockdown

**Appendix Figure 3:**
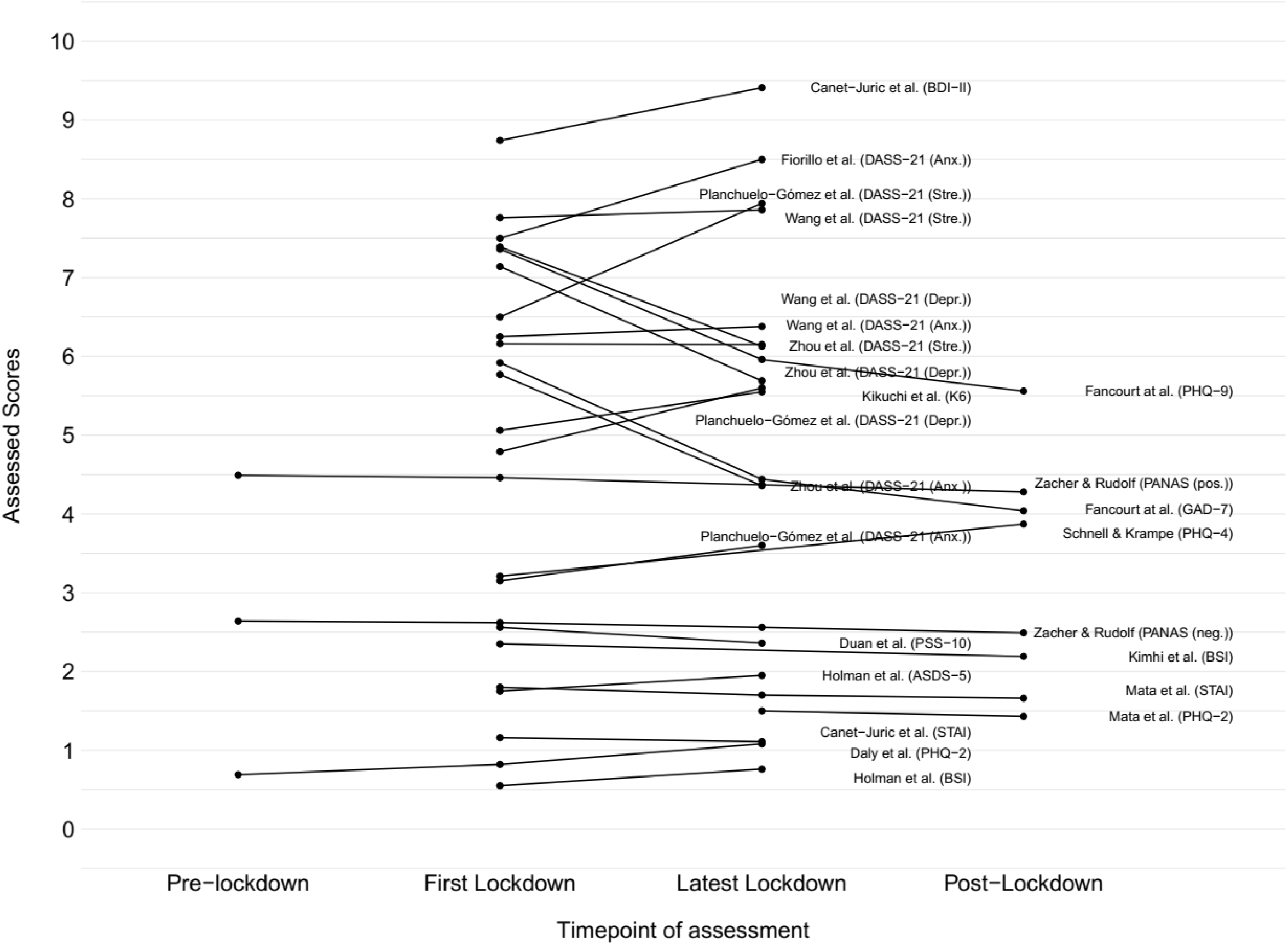
Low scores (< 10) of mental health problem assessment instruments during and after lockdown

**Appendix Figure 4:**
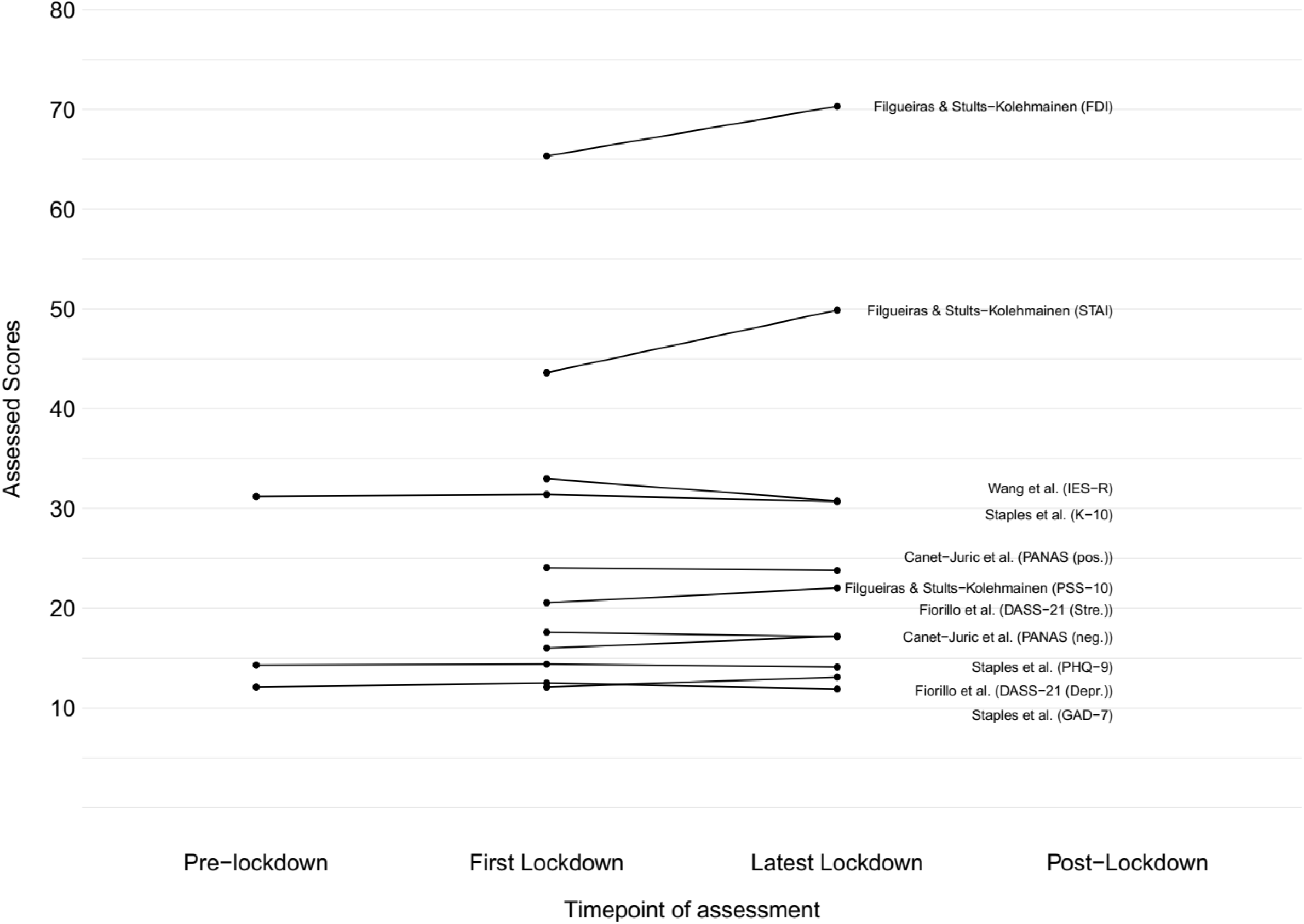
High scores (> 10) of mental health problem assessment instruments during and after lockdown

**Caption Figure 2 appendix:** DASS-21 (Anx.), Depression, Anxiety, Stress Scale - Anxiety; DASS-21 (Depr.), Depression, Anxiety, Stress Scale - Depression; DASS- 21 (Stre.), Depression, Anxiety, Stress Scale - Stress; FDI, Filgueiras Depression Inventory; GAD-2, Generalised Anxiety Disorder-2; GAD-7, Generalised Anxiety Disorder-7; GHQ-12, 12-item General Health Questionnaire; HADS (Anx.), Hospital Anxiety and Depression Scale; HADS (Depr.), Hospital Anxiety and Depression Scale; K6, Kessler Psychological Distress Scale; PHQ-2, Patient Health Questionnaire-2; PHQ-4, Patient Health Questionnaire-4; PHQ-9, Patient Health Questionnaire-9; PSS-10, Perceived Stress Scale-10; STAI, State-Trait Anxiety Inventory

**Caption Figure 3 appendix:** ASDS-5, Acute Stress Disorder Scale-5; BDI-II, Beck Depression Inventory-II; BSI, Brief Symptom Inventory; DASS-21 (Anx.), Depression, Anxiety, Stress Scale - Anxiety; DASS-21 (Depr.), Depression, Anxiety, Stress Scale - Depression; DASS-21 (Stre.), Depression, Anxiety, Stress Scale - Stress; GAD-7, Generalised Anxiety Disorder-7; K6, Kessler Psychological Distress Scale; PANAS (neg.), Positive and Negative Affect Schedule -Negative Affect; PANAS (pos.), Positive and Negative Affect Schedule -Positive Affect; PHQ-2, Patient Health Questionnaire-2; PHQ-4, Patient Health Questionnaire-4; PHQ-9, Patient Health Questionnaire-9; PSS-10, Perceived Stress Scale-10; STAI, State-Trait Anxiety Inventory

**Caption Figure 4 appendix:** DASS-21 (Depr.), Depression, Anxiety, Stress Scale - Depression; DASS-21 (Stre.), Depression, Anxiety, Stress Scale - Stress; FDI, Filgueiras Depression Inventory; GAD-7, Generalized Anxiety Disorder-7; IES-R, Impact of Event Scale-Revised; K-10, Kessler Psychological Distress Scale; PANAS (neg.), Positive and Negative Affect Schedule -Negative Affect; PANAS (pos.), Positive and Negative Affect Schedule -Positive Affect; PHQ-9, Patient Health Questionnaire-9; PSS-10, Perceived Stress Scale-10; STAI, State-Trait Anxiety Inventory

